# Inequalities in life expectancy in Australia according to education level: A whole-of-population record linkage study

**DOI:** 10.1101/2021.03.26.21254291

**Authors:** J Welsh, K Bishop, H Booth, D Butler, M Gourley, HD Law, E Banks, V Canudas Romo, RJ Korda

**Affiliations:** Research School of Population Health, Australian National University; School of Demography, Australian National University; Australian Institute of Health and Welfare

**Keywords:** life expectancy, socioeconomic, education, inequalities, mortality, record linkage

## Abstract

**Objective:** Life expectancy in Australia is amongst the highest globally, but national estimates mask within-country inequalities. We estimate education-related inequalities in adult life expectancy in Australia.

**Design and setting:** We estimated age-sex-education specific mortality rates using data from 2016 Australian Census linked to 2016-17 Death Registrations and standard life table methodology to calculate life expectancy.

**Participants:** 14,565,910 Australian residents aged 25 years and older.

**Main outcome measure:** Absolute (in years) and relative (ratios) differences in life expectancy at ages 25, 45, 65 and 85 years by sex and education level (5 levels, from Bachelor’s degree or higher [highest] to no secondary or post-secondary qualification [lowest]).

**Results:** At each age, those with lower education had shorter life expectancies. For men, the gap (highest vs lowest education) was 9.1 (95%CI: 8.8, 9.4) years at age 25, 7.3 (7.1, 7.5) years at age 45, 4.9 (4.7, 5.1) years at age 65 and 1.9 (1.8, 2.1) years at age 85. Absolute gaps were smaller for women compared with men but remained substantial: 5.5 (5.1, 5.9) years at age 25, 4.7 (4.4, 5.0) years at age 45, 3.3 (3.1, 3.5) years at 65 and 1.6 (1.4, 1.8) years at age 85. Relative differences were larger for men and increased with age.

**Conclusion:** Education-related inequalities in life expectancy from age 25 years in Australia are substantial such that those with the lowest education have a life expectancy equivalent to the national average 15-20 years ago. These vast gaps indicate large potential for further population health gains.

The known
Life expectancy in Australia has increased over time and is now among the highest in the world, but national estimates mask within-country inequalities and further potential for health gain. Area-level socioeconomic inequalities in health are well documented in Australia, but life expectancy gaps by individual-level socioeconomic position (e.g. highest level of education), a powerful measure of health inequality, have not been routinely available.

The new
Newly-available national linked data – 2016 Australian Census linked to Death Registrations (2016-17) – show that at each age (from age 25 years), men and women with lower levels of education, have shorter life expectancies. At age 25 years, the life expectancy gap between those with no educational qualifications and those with a university degree was 9.1 years for men and 5.5 years for women. Given lower life expectancy among men than among women, this represents a much larger relative – as well as absolute – gap for men.

The implications
Education-related gaps in life expectancy are considerable, larger than the OECD average, and mean that those with the lowest education have a life expectancy equivalent to the national average 15-20 years ago. The gaps indicate large potential for further improvements in population health, and health equity.

## Introduction

Life expectancy is a key metric for population health, and on this measure, Australia is one of the healthiest countries in the world [1]. This summary measure refers to the average number of additional years at a given age that survivors to that age are expected to live, given current mortality rates [2]. International comparisons of life expectancy are useful as they provide an indication of a country’s health progress and potential for health gain, and hence are routinely reported and used for monitoring purposes [3].

Cross-country comparisons in life expectancy, however useful, mask within-country differences. Estimates of life expectancy gaps between groups within a country are also needed; these estimates are not only powerful measures for highlighting inequalities within a country, they also provide valuable information on the potential for health gain from improvements in health equity, which is obscured in between-country international comparisons. Despite this, with the exception of Indigenous life expectancy gaps, between-population group inequalities in life expectancy are not commonly reported in Australia.

One of the most prominent forms of health inequality relates to an individual’s socioeconomic position, with well documented inequalities evident in numerous health indicators [4,5], including mortality [6,7]. Yet, there are no contemporary estimates of socioeconomic gaps in life expectancy for Australia. In this study, we use whole-of-population linked data to estimate inequalities in life expectancy in relation to education, an individual level measure of socioeconomic position, for men and women in Australia.

## Methods

### Data sources and study population

We used de-identified 2016 Census of Population and Housing data linked to Death Registrations (2016-17), available through the Multi-Agency Data Integration Project (MADIP). The Australian Bureau of Statistics link the data via the Person Linkage Spine, a person-level linkage key created by linking information from Medicare Enrolments, Social Security and Related Information, and Personal Income Tax, with virtually complete coverage of residents of Australia [8].

The scope of the 2016 Census was usual residents of Australia on 9 August 2016 [8] and the estimated person response rate was 94.8% [9]. Death Registrations available as part of the MADIP included all deaths registered in Australia [14] and were complete until the end of August 2017.

Our study sample included Australian residents aged at least 25 years at the time of the 2016 Census, or who would turn 25 years during the follow-up period, with a census record linked to the Spine.

### Variables

We derived highest level of education level from two self-reported census variables: highest year of secondary school completed and highest non-school qualification. We created five mutually exclusive categories: Bachelor’s degree or higher, irrespective of whether Year 12 was completed (highest education level); other post-secondary school qualification and completed Year 12; other post-secondary school qualification but did not complete Year 12; no post-secondary school qualification but completed Year 12; and, no post-secondary school qualification and did not complete Year 12 (lowest education level). Missing data on education level (6.2%, n=902,246) were imputed using single imputation by an ordered logistic model (see supplementary material for more information).

We obtained sex (male and female) from the Census, month and year of birth from the MADIP Spine, and month and year of death from Death Registrations. Details on how we derived day of birth and day of death (used to calculate exact age and follow-up time) are provided in supplementary material.

### Analysis

We estimated sex- and education-specific life expectancies (and 95% confidence intervals) at ages 25, 45, 65 and 85 years using standard life table procedures [2,10], using morality rates estimated from the linked Census-Deaths Registrations data. For each sex- and education-group, we estimated mortality rates by 5-year age group, starting from 25-29 years through to 100 years and older. We did this by applying education-specific rate ratios, derived from the Census linked to Death Registrations, to population mortality rates based on all deaths occurring in 2016 (see supplementary material). Individuals contributed person years-at-risk from the day they entered the study on 10 August, or when they turned 25 years, and were followed until day of death or the end of the study period on 31 August 2017, whichever occurred first. We accounted for birthdays over the study period and apportioned person-years by age. Time survived for individuals who died was the difference between date of death and date of entry into the study, aggregated by 5-year age group.

We estimated absolute inequalities in life expectancy as the difference, in years, between life expectancy for each education level and those with the highest education level. We estimated relative inequalities as a ratio of life expectancy between each education level and those with those with the highest education. For reporting purposes, we focus on the difference in life expectancy between those with the highest (Bachelor’s degree or higher) and lowest (no qualifications) education.

To facilitate comparison with OECD estimates, we also present education-related inequalities in life expectancy at ages 30 and 65 using a three-category measure of education: high education (Bachelor’s degree or higher, International Standard Classification of Education [ISCED levels 6-8]), intermediate education (secondary graduation with/without other non-tertiary qualifications, ISCED levels 3-5) and low education (no secondary school graduation or other qualification, ISCED levels 0-2).

We validated our estimates of life expectancy by conducting broad comparisons with official estimates of life expectancy at ages 25, 45, 65 and 85 years for men and women and in relation to an area-level measure of socioeconomic position at age 65 years (see supplementary material).

Analyses were conducted through the ABS virtual DataLab using Stata 16 [11] and R Studio [12].

## Results

There were 16,129,252 census records for Australian residents aged 25 years and older during the follow-up period. After excluding records that did not link to the Spine (n=1,562,623, 9.7%) or that linked in error (n=656, <0.01%), there were 14,565,910 persons in our sample. Among this sample, 140,954 deaths occurred in the follow-up period (98% of Death Registrations where age at death was ≥25 years linked to the Spine).

After imputation of missing education data, 26.8% (n=3,896,201) of the study population were classified as having a Bachelor’s degree or higher, 17.0% (n=2,474,557) as other post-secondary and Year 12, 15.2% (n=2,210,066) other post-secondary and no year 12, 12.5% (n=1,822,483) no post-secondary and Year 12 and 28.6% (4,162,603) no post-secondary and no Year 12. Proportions with high levels of education decreased with age (see Supplementary Table 1).

### Inequalities in life expectancy

Australian men and women with higher levels of education had longer life expectancies compared to those with lower levels of education (Table 1 and Supplementary Table 4). Life expectancy at age 25 was 61.2 years (95%CI: 61.0, 61.4) for men with a university degree compared with 52.1 years (51.9, 52.3) for men with no qualifications, a life expectancy gap of 9.1 (8.8, 9.4) years (Table 1). The male life expectancy gap was 7.3 (7.1, 7.5) years at age 45, 4.9 (4.7, 5.1) years at age 65 and 1.9 (1.8, 2.1) years at age 85.

**Table 1.** Absolute and relative differences by education level in life expectancy at ages 25, 45, 65 and 85 years for Australian men and women, 2016.

|  | Men<br>Life expectancy<br>(95%CI) | Difference<br>in years (95%CI) | Ratios | Women<br>Life expectancy<br>(95%CI) | Difference<br>in years (95%CI) | Ratios |
| --- | --- | --- | --- | --- | --- | --- |
| **Life expectancy by education level** |  |  |  |  |  |  |
| **At age 25 years** |  |  |  |  |  |  |
| Bachelor's degree (highest) | 61.2 (61.0, 61.4 ) | 0.0 | 1.00 | 63.6 (63.4, 63.9) | 0.0 | 1.00 |
| Other post-secondary & Yr12 | 59.5 (59.3, 59.7) | 1.7 (1.4, 2.0) | 0.97 | 63.3 (63.0, 63.5) | 0.3 (-0.1, 0.7) | 1.00 |
| Other post-secondary, no Yr12 | 57.6 (57.4, 57.7) | 3.6 (3.3, 3.9) | 0.94 | 61.9 (61.6, 62.1) | 1.7 (1.3, 2.1) | 0.97 |
| No post-secondary & Yr12 | 57.2 (56.9, 57.4) | 4.0 (3.6, 4.4) | 0.93 | 61.3 (61.1, 61.6 ) | 2.3 (1.9, 2.7) | 0.96 |
| No post-secondary, no Yr12 (lowest) | 52.1 (51.9, 52.3) | 9.1 (8.8, 9.4) | 0.85 | 58.1 (58.0, 58.3) | 5.5 (5.1, 5.9) | 0.91 |
| **At age 45 years** |  |  |  |  |  |  |
| Bachelor's degree (highest) | 41.6 (41.4, 41.8) | 0.0 | 1.00 | 44.0 (43.7, 44.2) | 0.0 | 1.00 |
| Other post-secondary & Yr12 | 40.3 (40.1, 40.5) | 1.3 (1.0, 1.6) | 0.97 | 43.8 (43.5, 44.0) | 0.2 (-0.2, 0.6) | 1.00 |
| Other post-secondary, no Yr12 | 38.6 (38.5, 38.8) | 3.0 (2.7, 3.3) | 0.93 | 42.7 (42.5, 42.9) | 1.3 (0.9, 1.7) | 0.97 |
| No post-secondary & Yr12 | 38.3 (38.0, 38.5) | 3.3 (2.9, 3.7) | 0.92 | 42.0 (41.8, 42.2) | 2.0 (1.6, 2.4) | 0.95 |
| No post-secondary, no Yr12 (lowest) | 34.3 (34.2, 34.4) | 7.3 (7.1, 7.5) | 0.82 | 39.3 (39.2, 39.4) | 4.7 (4.4, 5.0) | 0.89 |
| **At age 65 years** |  |  |  |  |  |  |
| Bachelor's degree (highest) | 22.9 (22.8, 23.1) | 0.0 | 1.00 | 25.0 (24.8, 25.2) | 0.0 | 1.00 |
| Other post-secondary & Yr12 | 22.1 (22.0, 22.3) | 0.8 (0.6, 1.0) | 0.97 | 25.0 (24.8, 25.3) | 0.0 (-0.4, 0.4) | 1.00 |
| Other post-secondary, no Yr12 | 20.9 (20.8, 21.0) | 2.0 (1.8, 2.2) | 0.91 | 24.2 (24.0, 24.4) | 0.8 (0.5, 1.1) | 0.97 |
| No post-secondary & Yr12 | 20.7 (20.5, 20.9) | 2.2 (1.9, 2.5) | 0.90 | 23.6 (23.4, 23.8) | 1.4 (1.1, 1.7) | 0.95 |
| No post-secondary, no Yr12 (lowest) | 18.0 (17.9, 18.1) | 4.9 (4.7, 5.1) | 0.79 | 21.7 (21.6, 21.8) | 3.3 (3.1, 3.5) | 0.89 |
| **At age 85 years** |  |  |  |  |  |  |
| Bachelor's degree (highest) | 7.64 (7.46, 7.83) | 0.0 | 1.00 | 8.67 (8.46, 8.88) | 0.0 | 1.00 |
| Other post-secondary & Yr12 | 7.87 (7.68, 8.07) | -0.2 (-0.5, 0.0) | 1.03 | 8.76 (8.52, 9.00) | -0.1 (-0.4, 0.2) | 1.01 |
| Other post-secondary, no Yr12 | 6.83 (6.72, 6.94) | 0.8 (0.6, 1.0) | 0.89 | 8.39 (8.21, 8.59) | 0.3 (0.0, 0.6) | 0.97 |
| No post-secondary & Yr12 | 7.40 (7.21, 7.60) | 0.2 (0.0, 0.5) | 0.97 | 8.45 (8.29, 8.60) | 0.2 (0.0, 0.5) | 0.97 |
| No post-secondary, no Yr12 (lowest) | 5.70 (5.64, 5.76) | 1.9 (1.8, 2.1) | 0.75 | 7.06 (7.02, 7.11) | 1.6 (1.4, 1.8) | 0.81 |

Note: Difference in life expectancy is the difference, in years, between each education group and the highest education level (Bachelor's degree). Life expectancy ratios is the ratio of life expectancy relative to the highest education level (Bachelor's degree).

For women, life expectancy at age 25 varied by up to 5.5 (5.1, 5.9) years between education levels, with life expectancy 63.6 years (63.4-63.9) for those with the highest education level and 58.1 years (58.0, 58.3) for those with the lowest level. The life expectancy gap between those with the highest and the lowest education was 4.7 (4.4, 5.0) years at age 45, 3.3 (3.1, 3.5) years at age 65 and 1.6 (1.4, 1.8) years at age 85.

Relative differences in life expectancy between education levels were larger for men than for women and generally larger with increasing age.

## Discussion

In Australia, life expectancy from age 25 years varies substantially according to level of education. Among men and women, life expectancy was lowest among those with no educational qualifications and rose with increasing education, at all ages. Absolute gaps between those with a university degree and those with no educational qualifications were substantial, as great as 9.1 years for men and 5.5 years for women aged 25 years, and decreased with age. Relative differences in life expectancy increased with age and were generally larger for men, reflecting shorter life expectancies.

Our study is the first to estimate education-related life expectancy with individual-level data with high population coverage and all of census linked prospectively to Death Registrations. Given this, there are few comparable studies with which to compare. However, we note that our estimates are larger than have been previously documented for Australia [3,13]. Australian data presented in an OECD report, based on death registrations linked to census data for 2011-12, found education-related gaps in life expectancy at age 25 years of 6.6 years for men and 3.7 years for women [3]. However, these were likely to be underestimated. The estimates were based on linked death registrations only (population denominators were sourced from external data sources) and the weights used to correct for the relatively low linkage rate (80%) did not account for differences in linkage rates by education [14], which are likely lower in lower education groups.

A degree of caution is required when directly comparing inequality estimates across countries, given that life expectancy estimates and the associated gaps can reflect differences in methodologies and data quality. However, our finding that education-related gaps in life expectancy are considerable is consistent with other international studies [3,15-17]. The average absolute highest - lowest education gap in life expectancy at age 25 years for 25 OECD countries, including Australia, based on linked census-mortality data for 2015-17, was 7.4 years for men and 4.5 years for women [3]. While our estimates exceed this by 1-2 years, they are comparable to estimates for males in Belgium (9.9 years) and Slovenia (8.3 years) and for females in Sweden (5.0 years), Denmark (5.1 years) and Hungary (5.7 years) [3].

In Australia, stark inequalities in life expectancy have been reported for other population groups. This includes a gap of 8.6 years in life expectancy at birth between non-Indigenous and Indigenous men and 7.8 years for women at age 25 years [1], and 5.0 and 4.0 years for men and women, respectively, living in rural and remote areas compared with those in major cities [18]. As expected, education-related inequalities based on individual data are larger than inequalities measured by area-level disadvantage (compare Table 1 [at age 65 years] and Supplementary Table 2; see also [4,19]). This is consistent with the fact that area-level measures are known to underestimate socioeconomic inequalities because of substantial within-area variation [20].

We found larger education-related differentials in life expectancy for men compared to women. This sex difference is typical of social differentials in general and has been shown to stem in part from the greater influence of men’s education and income on household resources, although this appears to be changing [21]. Our finding that life expectancy gaps are larger for men than women is consistent with previous research which has shown larger absolute and relative education-related inequalities in mortality rates for men compared to women, particularly from deaths caused by injuries and accidents and causes associated with smoking and alcohol use [6].

Australia performs well on life expectancy, which has been rising steadily and is among the highest in the world [1]. However, the findings from this study and elsewhere [13,22] highlight substantial inequalities in progress. The life expectancy reported here at age 25 for men with the lowest education in 2016 is equivalent to that for all males at age 25 in 1998, while for women the equivalent year is 2001 (58.3 years) [23], demonstrating a 15-20-year lag. Our findings highlight the potential for further substantial population health gains if socioeconomic inequalities were addressed.

The health system will play an important role in realising further population health gains. Previous Australian research found the largest absolute education-related inequalities were in chronic diseases amenable to prevention, including lung cancer, ischaemic heart disease and chronic lower respiratory disease [6]. Achieving reductions in health inequalities will also require addressing upstream social, economic and cultural determinants of health [24]. Ongoing monitoring of within-country inequalities in life expectancy can be used to assess the impact of policies on upstream determinants on health equity [25]. While data linkage quality may limit historical assessment of policies, data from the MADIP, assuming it is routinely updated, can facilitate ongoing monitoring.

## Strengths and limitations

In this record linkage study, life expectancy estimates were based on data with high coverage of the Australian population and virtually complete ascertainment of deaths. Education-specific mortality rates were estimated by applying education-specific rate ratios to mortality rates from the full population. This method assumes that the rate ratios estimated from the analysis file are unbiased and can be generalised to the full population. Although we had virtually complete (98%) ascertainment of deaths, a previous study using these data found evidence that among young women, socioeconomic inequalities in mortality rates were underestimated [6]. Given this, it is likely that our estimates also underestimate the true education-related gaps in life expectancy for women. Finally, estimates presented here should be interpreted as summaries of inequalities in life expectancy, given that the meaning of education (and its relationship to health) has cohort effects and that all life expectancy estimates are based on a hypothetical population ageing through mortality rates observed at one point in time.

## Conclusion

In Australia, education-related gaps in life expectancy are considerable, such that those with the lowest levels of education in 2016 had a life expectancy equivalent to the national average 15-20 years prior. They are larger than have been documented in many other OECD countries and highlight the substantial potential for further improvements in population health if socioeconomic inequalities in health were addressed.

## Supporting information

see supplementary material

## Data Availability

Data part of the Multi-Agency Data Integration Project are available for approved projects to approved government and non-government users.

https://www.abs.gov.au/websitedbs/D3310114.nsf/home/Statistical+Data+Integration+-+MADIP

## Ethics statement

Ethics approval for this study was granted by the Australian National University Human Research Ethics Committee (reference 2016/666).

## Role statement

JW and RK had full access to the data and conceived the study; JW, KB, HB, VCR and RK designed the methodological strategy; JW did the analytical calculations; JW wrote the first draft of the manuscript with advice from KB, HB, VCR, DB, HDL, MG, EB and RK. All authors provided critical feedback and approved the final manuscript.

## Data sharing statement

Data part of the Multi-Agency Data Integration Project are available for approved projects to approved government and non-government users. https://www.abs.gov.au/websitedbs/D3310114.nsf/home/Statistical+Data+Integration+-+MADIP

## Acknowledgements

We acknowledge the contributions of members of the Whole-of Population Linked Data Project team, including Chief Investigators: Walter Abhayaratna, Nicholas Biddle, Bianca Calabria, Louisa Jorm, Raymond Lovett, John Lynch and Naomi Priest; Associate Investigators: Tony Blakely and Rosemary Knight; Partner Investigators: James Eynstone-Hinkins, Louise Gates, Michelle Gourley, Gary Jennings, Lynelle Moon, Lauren Moran, Talei Parker, Clare Saunders and Bill Stavreski; and Project Manager: Katie Beckwith. The authors also wish to thank Grace Joshy.

