## Supplementary material for "Inequalities in life expectancy in Australia according to education level: A whole-of-population record linkage study": see supplementary material

**Manuscript title**

**Linked 2016 Census and Death Registrations**

We used de-identified 2016 Census of Population and Housing and Death Registrations (2016-17), linked through the Multi-Agency Data Integration Project (MADIP). Underpinning all MADIP data is the Person Linkage Spine. The Spine creates a person-level linkage key, with virtually complete coverage of residents of Australia, created by linking information from three administrative databases: Medicare Enrolments, Social Security and Related Information, and Personal Income Tax [1]. The Australian Bureau of Statistics link all data within the MADIP via the Spine. Census records were linked to the Spine using deterministic and probabilistic linking methods, on name, full date of birth, address and sex, achieving a linkage rate of 92% [1]. Death Registrations were linked to the Spine using deterministic linking methods with a linkage rate of 97%.

**Estimating person-years-at-risk**

The available linked census and deaths data included month and year of birth and month and year of death, but not day.

*Calculating day of birth:* All births were assumed to have occurred on the 15th day of the month, except for those who died in their birth month; for a more accurate approximation of person-years-at-risk among the latter group, their births were assumed to have occurred on the 1st of the month.

*Calculating day of death:* Deaths were assumed to have occurred on the 15th of the month, with two exceptions to derive more accurate approximations of person-years-at-risk. First, deaths that occurred in August 2016 were assumed to have occurred on the 21st of the month (the mid-point of the follow-up time for that month). Second, among those who died in their birth month, deaths were assumed to have occurred on the 10th of the month for those who died at census age, and on the 22nd of the month for those who died at census age+1. Age at death was available in the Death Registration data.

**Imputation of missing education data**

Missing data on education level (6.2%, n=902,246) were imputed using single imputation by an ordered logistic model with age, sex, broad cause of death and a set of auxiliary variables as predictors. The auxiliary variables, chosen because they were associated with missingness or correlated with the 5-level education variable, included: Socio-Economic Indexes for Areas (SEIFA) Index of Education and Occupation measured at the Statistical Area level 2 (SA2) in population-based quintiles [2], personal income category, occupation, engagement in employment, training or education, English language proficiency, work hours, dwelling type, internet access at home and place of usual residence one year ago.

**Method for estimating education-specific mortality rates**

Previous validation of 2016 Census linked to Death Registration (2016-17) data has shown that mortality rates in the linked file differ to those in the Australian population: mortality rates in the linked file were lower at younger ages and slightly higher at older ages compared to rates estimated using the complete Death Registrations file, see [3]. If left uncorrected, this would result in an underestimate of mortality rates in the linked Census-Death Registration file, ultimately overestimating life expectancy.

To correct for differences in mortality rates in the linked Census to Death Registrations, we estimated education-specific mortality rate ratios (deaths/ person-years) from the linked analysis file using negative binomial regression, and population mortality rates (deaths in 2016/ 2016 mid-year estimated resident population) using Poisson regression and deaths taken from the complete Death Registrations file. We then estimated education-specific mortality rates by applying education specific rate ratios to population mortality rates.

**Validation analyses**

*Methods:* Before investigating inequalities in life expectancy, we validated our estimates by conducting broad comparisons of our estimates with available estimates produced by the Australian Bureau of Statistics. These included estimates from 2014-16 of sex-specific life expectancy at ages 25, 45 and 65 [4], and estimates of life expectancy at age 65 in relation to an area-level measure of socioeconomic position. The area-level measure of socioeconomic position was the SEIFA Index of Relative Socio-Economic Disadvantage (IRSD), measured at the SA2 level in population-based quintiles [5].

*Results:* Our estimates of life expectancy derived from 2016 population mortality rates by sex and IRSD quintile, were greater than, but within 0.4 years, of official estimates for 2014-16 (Supplementary Table 2). Comparison of estimates of life expectancy at age 65 years by IRSD quintile derived from our linked analysis file compared to official estimates revealed small differences between our estimates and official estimates in relation to IRSD quintile (ratio of differences ranged from 0.98 to 1.05 for men and 1.00 to 1.03 for women, Supplementary Table 3).

**Supplementary Tables**

Supplementary Table 1. Proportion (%) of the Australian population with each education level by broad age group and sex, 2016.

|  | Men | Women |
| --- | --- | --- |
| **25-44 years** |  |  |
| Bachelor’s degree (highest) | 32.3 | 42.4 |
| Other post-secondary & Yr12 | 24.1 | 22.2 |
| Other post-secondary, no Yr12 | 14.7 | 8.9 |
| No post-secondary & Yr12 | 15.7 | 15.6 |
| No post-secondary, no Yr12 (lowest) | 13.2 | 11.0 |
| **45-64 years** |  |  |
| Bachelor’s degree (highest) | 23.0 | 25.6 |
| Other post-secondary & Yr12 | 15.6 | 14.7 |
| Other post-secondary, no Yr12 | 25.1 | 15.9 |
| No post-secondary & Yr12 | 11.6 | 13.0 |
| No post-secondary, no Yr12 (lowest) | 24.7 | 30.8 |
| **65-84 years** |  |  |
| Bachelor’s degree (highest) | 15.9 | 13.7 |
| Other post-secondary & Yr12 | 12.3 | 8.0 |
| Other post-secondary, no Yr12 | 24.5 | 10.3 |
| No post-secondary & Yr12 | 10.6 | 11.5 |
| No post-secondary, no Yr12 (lowest) | 36.6 | 56.5 |
| **85 years and older** |  |  |
| Bachelor’s degree (highest) | 10.4 | 5.7 |
| Other post-secondary & Yr12 | 10.2 | 4.5 |
| Other post-secondary, no Yr12 | 20.9 | 6.5 |
| No post-secondary & Yr12 | 10.2 | 11.1 |
| No post-secondary, no Yr12 (lowest) | 48.4 | 72.2 |

Notes: Estimates are taken from ABS TableBuilder using 2016 Census. Australian residents with missing education have been excluded.

Supplementary Table 2. Comparison of estimated life expectancy at ages 25, 45, 65 and 85 years for Australian men and women with official estimates produced by the Australian Bureau of Statistics (ABS)

|  | Men | | | Women | | |
| --- | --- | --- | --- | --- | --- | --- |
|  | Analysis file  2016-17 | ABS estimate  2015-17 | Difference  (years) | Analysis file  2016-17 | ABS estimate  2015-17 | Difference  (years) |
| 25 years | 56.6 (56.1, 56.6) | 56.2 | 0.4 | 60.4 (60.3, 60.5) | 60.1 | 0.3 |
| 45 years | 37.5 (37.4, 37.6) | 37.2 | 0.3 | 40.9 (40.9, 41.0) | 40.7 | 0.2 |
| 65 years | 19.9 (19.8, 20.0) | 19.7 | 0.2 | 22.5 (22.5, 22.6) | 22.3 | 0.2 |
| 85 years | 6.41 (6.36, 6.46) | 6.26 | 0.2 | 7.41 (7.37, 7.45) | 7.30 | 0.1 |

Note: ABS estimates are taken from: Australian Bureau of Statistics. Life Tables, States, Territories and Australia, 2015–2017. Cat. no. 3302.0.55.001. Canberra: ABS; 2018.

Supplementary Table 3. Comparison of estimated life expectancy at age 65 years for Australian men and women by SEIFA IRSD with official estimates produced by the Australian Bureau of Statistics (ABS)

|  | Men | | | | Women | | | | |
| --- | --- | --- | --- | --- | --- | --- | --- | --- | --- |
|  | Life expectancy | |  |  | | Life expectancy | |  |  |
|  | Analysis file, 2016-17 | ABS estimates 2014-16 | Difference  in years | Ratios | | Analysis file, 2016-17 | ABS estimates 2014-16 | Difference  (years) | Ratio |
| Most disadvantaged | 18.6 | 18.1 | 0.5 | 1.03 | | 21.7 | 21.2 | 0.5 | 1.02 |
| 2 | 19.5 | 18.6 | 0.9 | 1.05 | | 22.1 | 21.6 | 0.5 | 1.02 |
| 3 | 20.1 | 19.5 | 0.6 | 1.03 | | 22.6 | 22.0 | 0.6 | 1.03 |
| 4 | 20.6 | 20.7 | -0.1 | 1.00 | | 23.2 | 22.9 | 0.3 | 1.01 |
| Least disadvantaged | 21.2 | 21.7 | -0.5 | 0.98 | | 23.5 | 23.6 | -0.1 | 1.00 |

Notes: SEIFA IRSD is Socio-Economic Indexes for Areas, Index of Relative Socio-economic Disadvantage, measured in population-based quintiles. ABS estimates are taken from: *Australian Burden of Disease Study: impact and causes of illness and death in Australia 2015*. Canberra: AIHW; 2015.

Supplementary Table 4. Absolute and relative inequalities in life expectancy by education at ages 30 and 65 for Australian men and women, 2016.

|  | Men | | | Women | | |
| --- | --- | --- | --- | --- | --- | --- |
|  | Life expectancy (95%CI) | Difference  in years (95%CI) | Ratios | Life expectancy  (95%CI) | Difference  in years (95%CI) | Ratios |
| Life expectancy by education level | |  |  |  |  |  |
| At age 30 years |  |  |  |  |  |  |
| High education | 56.3 (56.1, 56.5) | -- | 1.00 | 58.7 (58.4, 58.9) | -- | 1.00 |
| Intermediate education | 53.2 (53.1, 53.3) | 3.1 (2.8, 3.3) | 0.95 | 57.2 (57.1, 57.4) | 1.4 (1.2, 1.7) | 0.98 |
| Low education | 47.5 (47.3, 47.6) | 8.8 (8.5, 9.1) | 0.84 | 53.3 (53.2, 53.4) | 5.4 (5.1, 5.6) | 0.91 |
| At age 65 years |  |  |  |  |  |  |
| High education | 22.9 (22.7, 23.1) | -- | 1.00 | 25.0 (24.8, 25.2) | -- | 1.00 |
| Intermediate education | 21.2 (21.1, 21.2) | 1.8 (1.5, 2.0) | 0.92 | 24.2 (24.0, 24.3) | 0.8 (0.5, 1.1) | 0.97 |
| Low education | 18.0 (17.9, 18.1) | 4.9 (4.7, 5.2) | 0.78 | 21.7 (21.6, 21.8) | 3.3 (3.0, 3.5) | 0.87 |

Note: Difference in life expectancy is the difference, in years, between each education group and the highest education level (Bachelor’s degree). Life expectancy ratios is the ratio of life expectancy relative to the highest education level (Bachelor’s degree). Education categories: High education refers to a Bachelor’s degree or higher (International Standard Classification of Education [ISCED] levels 6-8), intermediate education refers to secondary school graduation with/without other non-tertiary qualifications (ISCED levels 3-5) and low education refers to no secondary school graduation or other qualification (ISCED levels 0-2).

5.	Australian Institute of Health and Welfare. Australian Burden of Disease Study: impact and causes of illness and death in Australia 2015. Canberra: AIHW; 2019.
